# Quantitative COVID-19 infectiousness estimate correlating with viral shedding and culturability suggests 68% pre-symptomatic transmissions

**DOI:** 10.1101/2020.05.07.20094789

**Authors:** Meher K. Prakash

## Abstract

A person clinically diagnosed with COVID 19 can infect others for several days before and after the onset of symptoms. At the epidemiological level, this information on how infectious someone is lies embedded implicitly in the serial interval data. Other clinical indicators of infectiousness based on the temporal kinetics of the viral shedding from the nasopharyngeal swabs and sputum show the former decaying weeks sooner than the latter. In this work, we attempt to provide a better quantitative estimate for the temporal infectiousness profile using serial interval data from a combined 1251 individuals reported in the literature. We show that the infectiousness profile which we calculate correlates well with the viral shedding kinetics from nasopharyngeal swabs (*ρ*=0.97, p=0.00) and culturability (*ρ*=0.83, p=0.01). The profile suggests that a 68.4% (95% CI: 67.0-69.7%) of the infections are caused by infections before the symptoms appear, which is a much stronger pre-symptomatic influence than what was predicted in the literature 44% (95% CI: 25-69%) using serial data from 77 individuals.

## Introduction

The epidemiological parameter R-naught (R_0_) signifies how many new infections are caused on average by an infected person. The R_0_ of COVID-19, for which some of the estimates have been around 2.2 to 2.5,^1,2^ is a regularly monitored parameter to see if the spread of the infection is contained. Going one step further into understanding the origins of R_0_, the spread of infections depends on the physical contact between the infected and the susceptible individuals, as well as on how long an infected person can infect others. A substantial amount of infections of COVID-19 are transmitted through asymptomatic individuals,^3^ and their infectiousness profiles are harder to track. Here we specifically focus on the data from the symptomatic individuals to quantify their temporal infectiousness profile during the course of their personal history of infection.

On the clinical side, the viral load of the SARS-CoV-2 in the nasopharyngeal swab, oropharyngeal swab, sputum, blood, urine and stool specimens of the COVID-19 patients can be estimated using PCR with reverse transcriptase (RT-PCR). The cycle threshold (C_t_) values from RT-PCR are converted to RNA copy numbers in the sample and interpreted. Several studies followed the temporal history of SARS-CoV-2 RNA shedding profiles in individuals for many weeks after the onset of symtpoms.^4-7^ Some of these studies showed that peak viral loads were higher for older individuals,^6^ whereas, others on a larger cohort showed no significant differences in viral shedding with sex, age group, and the severity of the disease.^7^

Despite this apparent commonality across the patient demographics, the viral shedding patterns from the different specimens show varying kinetics. While some reports show that the viral load from the nasal swabs showed much faster decay within about a week after the symptoms, compared to throat swabs,^4^ other reports found no discernible differences.^8^ Further, using sputum or stool analysis a significant number of patients are considered positive for 3 to 4 weeks,^8,9^ but this period is about 1 to 2 weeks using a serum or swab analysis.^8,9^ The primary reason for the apparent conflict comes from the fact that the RNA load inferred from RT-PCR does not clarify what fraction of it is culturable or infectious.

To develop insights into the temporal infectiousness profile, which is important from the public health point of view, one must know which data to focus on. In this sense, it is important to juxtapose the clinically measured viral loads against epidemiological data to find a consistent interpretation. The serial interval data, which is the time between the infectee and the infected showing clinical symptoms, from 77 patient histories was used to infer the infectiousness profile.^7^ The viral load data was not used to predict this serial interval, and no direct comparison of the inferred infectiousness profile with the viral shedding was made.^7^

In this work, we combine the serial interval data of 1251 individuals from three different works: 689 patients from Ma et al.,^11^ 485 patients from Du et al.,^10^ and 77 patients from He et al.^7^, to form a larger dataset that is likely to capture the underlying distribution better.

## Results and Discussion

Epidemiologically, serial interval is constructed from knowing the days when the symptoms appear in the infectee and the infected. To estimate it, however, one should implicitly know the incubation times (*f*) of both as well as the infectiousness or the transmissibility of the infection (*g*) on the day infectee transmits to the infected (**Supplementary Figure 1**). Using the data from 181 patients, the incubation time distribution was approximated^12^ a Weibull distribution with parameters *λ*=6.258, *k*=2.453 and other reports^13^ agree with this estimate. We thus use the serial interval data from the 1251 individuals, and this incubation estimate to solve the inverse problem of estimating the infectiousness profile, starting from the day an individual is infected (**Methods**). The criterion used for accepting the estimate was that it maximizes the R^2^ between the predicted and the observed serial interval. The best estimate for the serial interval is shown in **Figure 1**, and the serial interval that was used for obtaining this prediction was a Weibull distribution with a median of 3.1 days and is shown in **Figure 2A**. The peak of infectiousness is at 2 days after the infection, and it decreases after it. The effect of the incubation period was averaged over, and the infectiousness profile relative to the day of the onset of the symptoms was obtained (**Figure 2B**). The infectiousness profile shows that a large part of the infectiousness (68.4%) finishes before the onset of the symptoms. The 95% confidence interval on this estimate was obtained using bootstrapping (**Methods**) as (67.0-69.7%). A sensitivity analysis performed with (*λ*=5.2,*k*=2.0) and (*λ*=7.0,*k*=1.5) resulted in consistent estimates for the presymptomatic infections: 65.04% (95% CI: 64.5-65.5%) and 69% (95% CI: 67.8-71.1%) respectively. The average value of the pre-symptomatic infectiousness is much higher than the prediction in the literature 44% (95% CI: 25-69%).^7^ The smaller variation is possibly from the larger data set we have used as well as their use of the incubation data from a smaller set of 10 out of the 425 patients studied by Li et al.^1^

**Figure 1.**
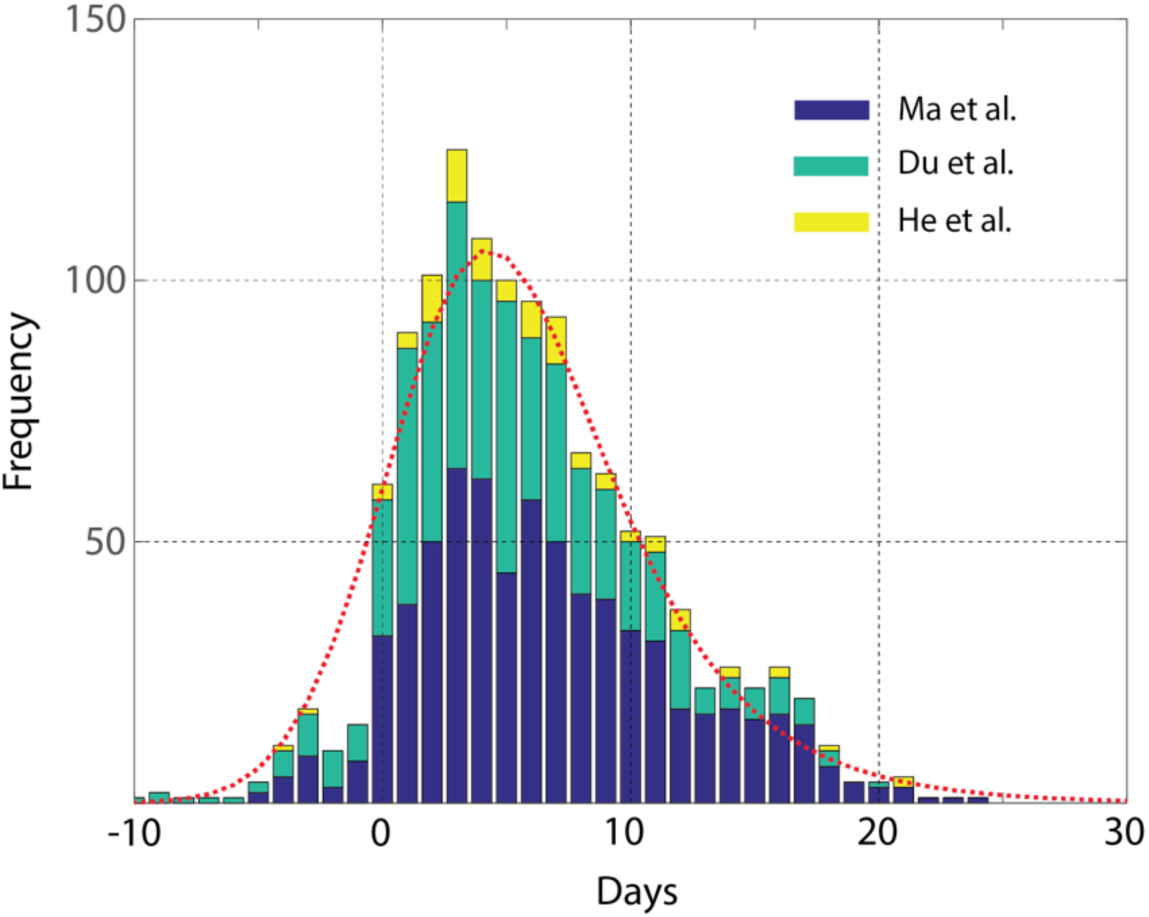
Inferring infectiousness from serial interval. We combined the serial interval data of 689 patients from Ma et al.^11^, 485 patients from Du et al.^10^, and 77 patients from He et al.,^7^ to build a larger data set. The inverse problem of the infectiousness profile was solved by assuming an incubation period from Lauer et al.^16^ The serial interval predicted using this estimate is shown in dotted red line. While Ma et al.^11^, Du et al.^10^, showed fits to their serial interval data, He et al.^7^ made a similar estimate for the infectiousness based on the smaller data size of 77 patients.

**Figure 2.**
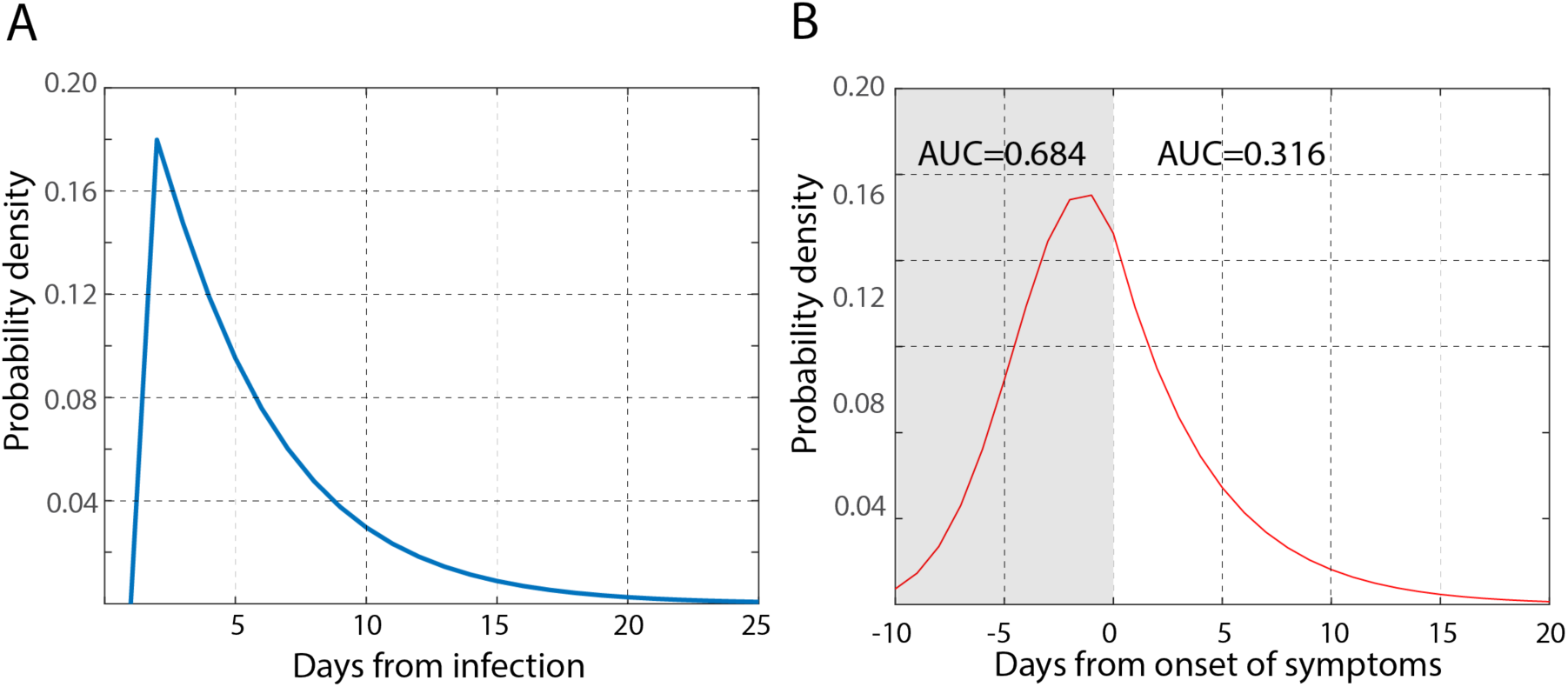
Infectiousness profiles. The infectiousness that was inferred from the serial interval data is shown in two different representations. We assumed that the infectiousness profile starting from the day someone is infected follows a Weibull distribution. Using the quality of the linear regression between the predicted serial interval frequencies and the observed ones from Figure 1 as the optimization criterion, we estimated the parameters to be *λσ*=4.4, *k*=1.04 over the parameter range. **A**. The infectiousness peaks 2 days after the start of the infection, **B**. and relative to the onset of symptoms, the infectiousness is 68.4% (95% CI: 67.069.7%) complete by the time the onset of symptoms begins.

In order to develop a comprehensive picture of the infectiousness, we compare the profile we estimated to the two clinical laboratory markers: the viral shedding kinetics and culturability. Such relations have been developed earlier for influenza^14^ and HIN1.^15,16^ **Figure 3** shows that the infectiousness profile we estimate in **Figure 2B** has similar kinetics to the viral shedding kinetics from the nasopharyngeal swab samples. Only the data from the longitudinal studies on individuals was used for this comparison. The chance of obtaining a positive culture decreases with time and its kinetics is also consistent with the viral shedding and infectiousness (**Figure 3**). In addition to this kinetics, we also study the correlations between the infectiousness we estimate and the average viral shedding (**Figure 4A**) and the chance of obtaining a positive culture (**Figure 4B**) that was calculated by combining data from two different works.^8,17^ Both these correlations are very strong, with correlation coefficients 0.83 and 0.97, respectively and high significance.

**Figure 3.**
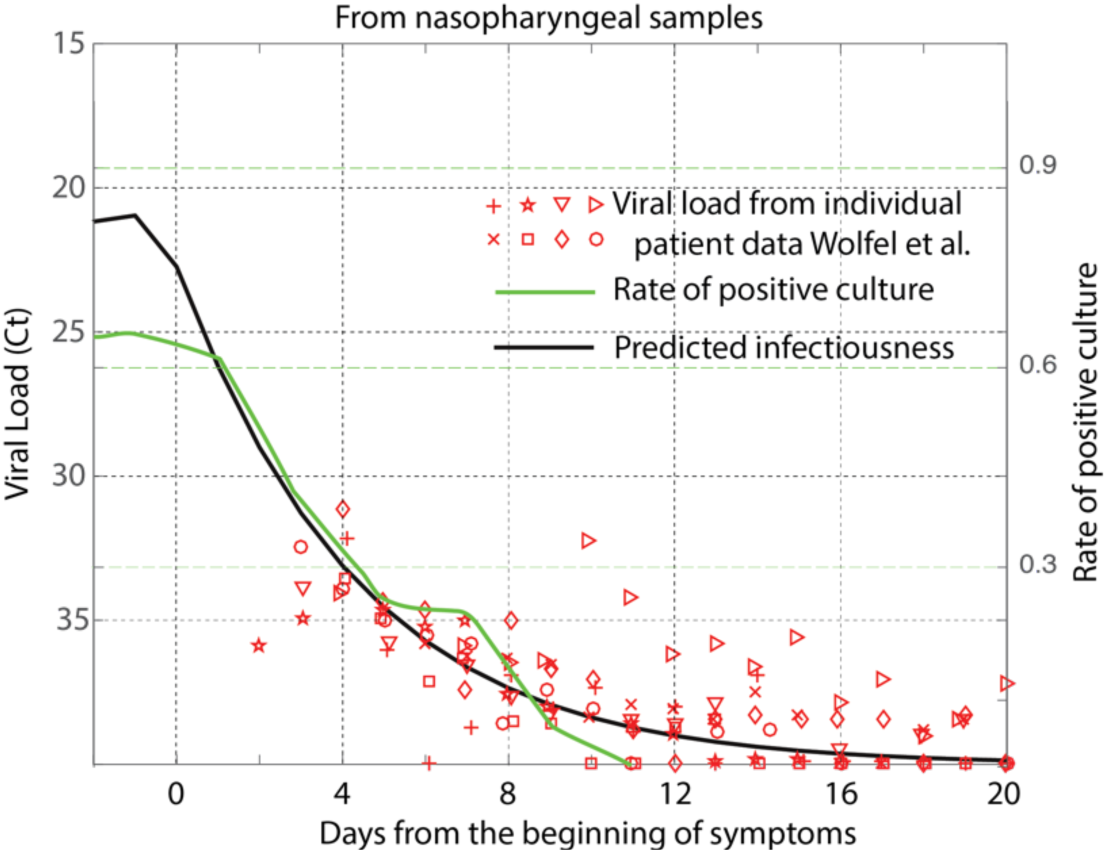
COVID-19 infectiousness kinetics. The predicted drop in infectiousness is compared to the viral shedding data measured as the cycle threshold (C_t_) in the RT-PCR, and the rate of positive culture. The longitudinal viral shedding data gathered using nasopharyngeal swabs for at least 20 days after the onset of the symptoms^8^ was used in this analysis. The rate of a positive culture, defined as the (number of positive cultures)/(number of positive + number of negative cultures) was obtained from the combined data on cultures from Wolfel *et al*.^8^ and Arons *et al*.^12^. Our predictions show similar kinetic trends, as the positive culture rate and the nasal swab samples, unlike the saliva samples (**Supplementary Figure 2**).

**Figure 4.**
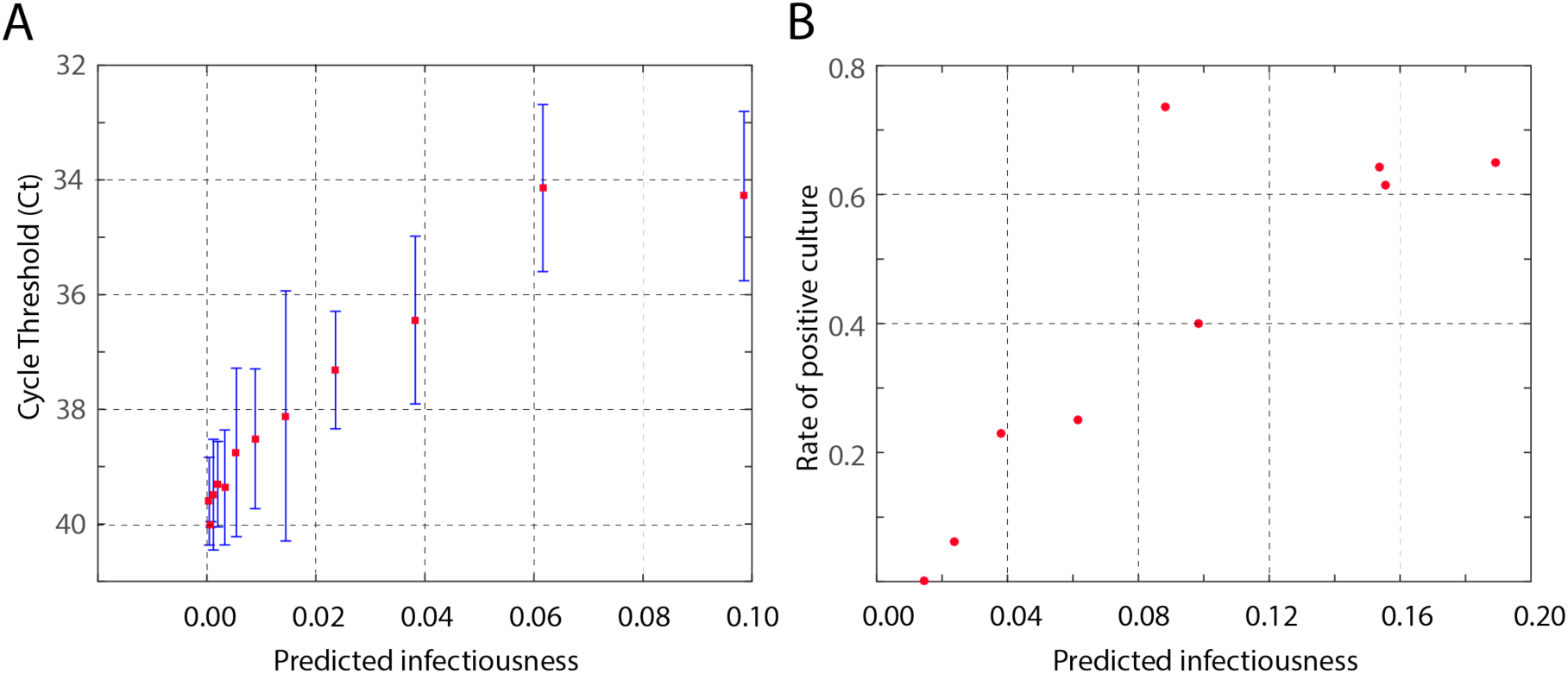
Correlating infectiousness. The predicted infectiousness is plotted relative to the A. Cycle threshold (C_t_) in the RT-PCR of nasal swab samples and B. rate of positive culture. The correlation coefficients are 0.83 and 0.97 respectively. A two-day binning of the data was used in this graphic. The longitudinal data of the viral shedding available from day 4,^7^ while the positive culture data was obtained from day 3,^7^ and day -6.^12^ The differences in the availability of the data is reflected in the range of infectiousness plotted in these two figures.

Clearly, the kinetics of the samples from sputum show a much slower decay (**Supplementary Figure 2**). Which means that if one uses the sputum samples as a criterion for understanding the infectiousness, the C_t_ threshold will be much higher, comparing both the observed culturability and the infectiousness we estimate.

## Conclusions

By using combined serial interval data from several sources, we could estimate the temporal infectiousness profile which suggests with high confidence that about 68% of the infections are likely to be caused before the onset of symptoms. The kinetics of the viral shedding, culturability and the infectiousness were all found to be consistent which suggests the possibility of using them as surrogates for one another.

## Methods

### Infectiousness estimation

The serial interval is illustrated in **Supplementary Figure 1**. The serial interval of (*τ* + *τ*_2_- *τ*_1_) days arises from infectiousness at *τ*, and the incubation times *τ*_1_, *τ*_2_ of the infectee and the infected. The serial interval distribution *h*(*t*) is given as

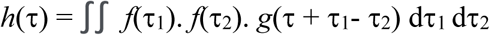

where *f(t)* is the incubation distribution and g(t) is the infectiousness profile *t* days after the infectee gets infected. *f(t)* and *g*(*t*) were both assumed to follow Weibull distributions. The parameters *λ*=6.258, *k*=2.453 were used for *f*(*t*). A 10 × 10 grid was used to scan through the parameters *λ*, *k* for *g*(*t*). Corresponding to each choice of the parameters, the predicted serial interval was plotted against the observed serial interval data. The R^2^ of the linear regression between the predictions and the observations was maximized and the *λ* and k were identified.

### Bootstrapping

To estimate the confidence intervals, bootstrapping with 1,000 replicates was used. From the serial interval data of 1251 individuals, 1000 independent sample sets each of size 300 were randomly selected by replacing the numbers each time. The calculations for infectiousness estimation were repeated as mentioned above with a 10 × 10 grid for scanning over *λ* and *k*. Using the best estimates for the serial interval in each replicate, the distribution of the infectiousness before symptoms was obtained. This distribution was used to obtain the 95% confidence interval.

## Data Availability

All codes and analysis shall be made available on request

## Supplementary Figure

**Supplementary Figure 1.**
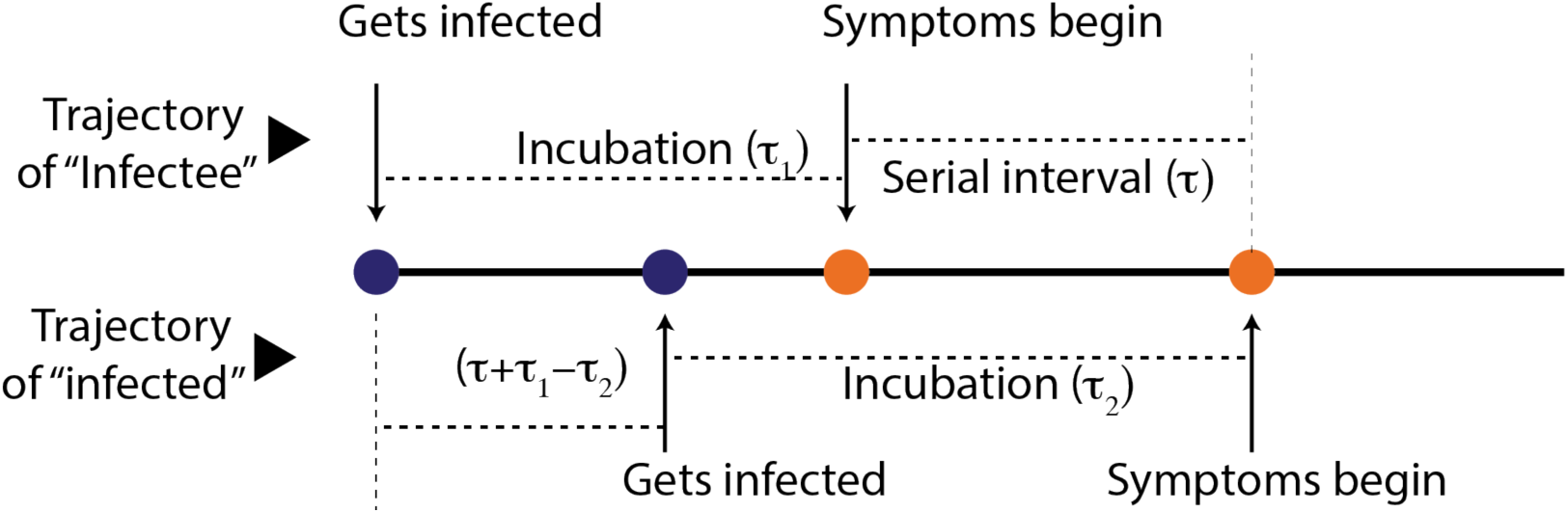
Serial interval. The time between the two appearance of symptoms in the infectee and the infected is illustrated. The infectiousness at *τ*, and the incubation times *τ*_1_, *τ*_2_ of the infectee and the infected are all combined to find the chance of a serial interval of (*τ* + *τ*_2_- *τ*_1_) days.

**Supplementary Figure 2.**
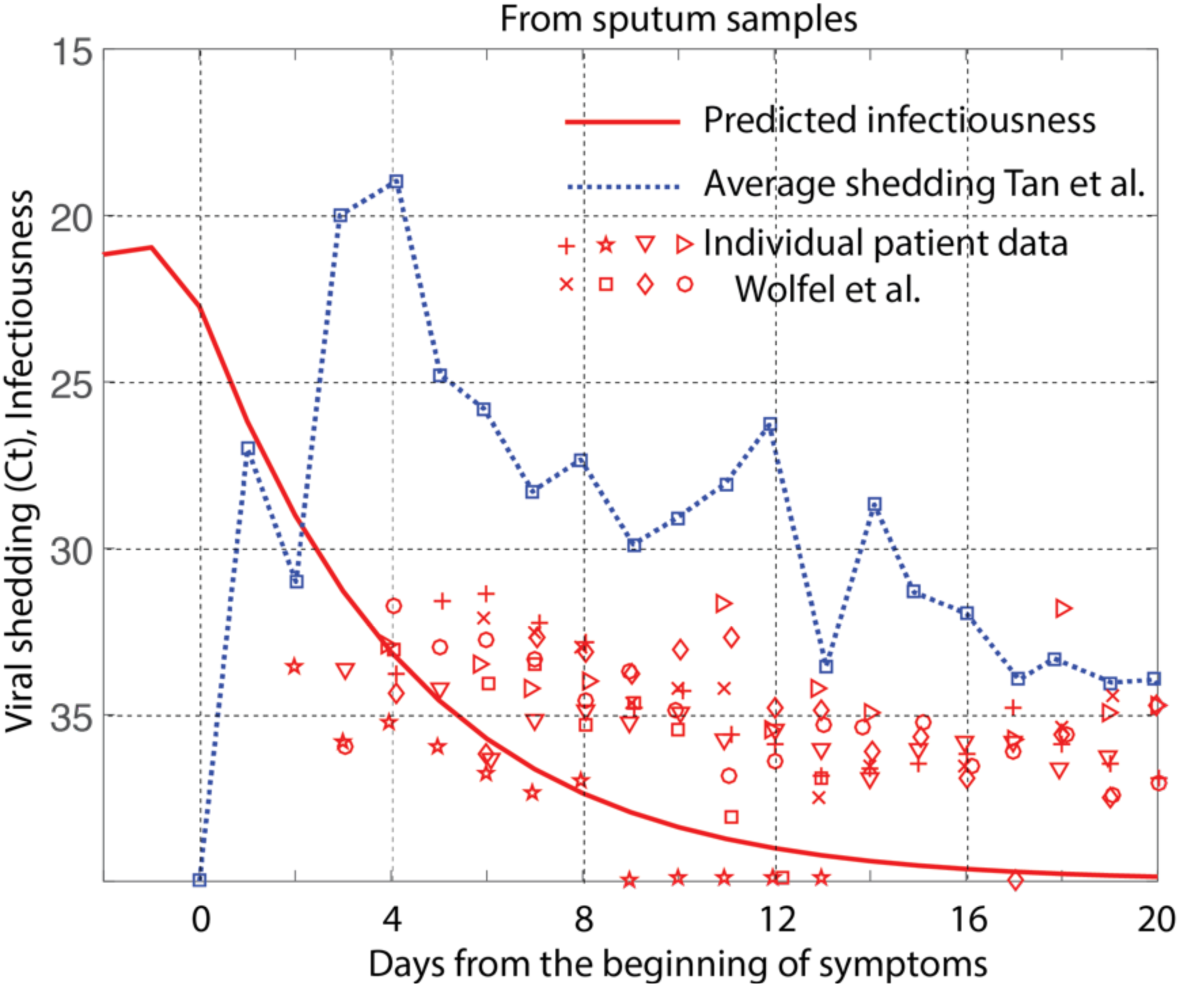
COVID-19 kinetics from sputum samples. The predicted drop in infectiousness as shown in Figure 3 is compared with the viral shedding data (C_t_) measured from the sputum samples. The longitudinal individual patient data from Wolfel et al.^8^ and the average profile from Tan et al.^9^ are compared.

